# Wearable-Derived Maternal Heart Rate Variability As A Novel Digital Biomarker of Preterm Birth

**DOI:** 10.1101/2022.11.04.22281959

**Authors:** Summer R. Jasinski, Shon Rowan, David M. Presby, Elizabeth A. Claydon, Emily R. Capodilupo

## Abstract

Despite considerable health consequences from preterm births, their incidence remains unchanged over recent decades, due partially to limited screening methods and limited use of extant methods. Wearable technology offers a novel, noninvasive, and acceptable way to track vital signs, such as maternal heart rate variability (mHRV). Previous research observed that mHRV declines throughout the first 33 weeks of gestation in term, singleton pregnancies, after which it improves. The aim of this study was to explore whether mHRV inflection is a feature of gestational age or an indication of time to delivery. This retrospective case-control study considered term and preterm deliveries. Remote data collection via non-invasive wearable technology enabled diverse participation with subjects representing 42 US states and 16 countries. Participants (N=241) were recruited from the WHOOP (Whoop, Inc.) userbase and wore WHOOP straps during singleton pregnancies between March 2021 and October 2022. Mixed effect spline models by gestational age and time until birth were fit for within-person mHRV, grouped into preterm and term births. For term pregnancies, gestational age (AIC = 26627.6, R^2^m = 0.0109, R^2^c= 0.8571) and weeks until birth (AIC =26616.3, R^2^m = 0.0112, R^2^c = 0.8576) were representative of mHRV trends, with significantly stronger fit for weeks until birth (relative log-likelihood ratio = 279.5). For preterm pregnancies, gestational age (AIC =1861.9, R^2^m = 0.0016, R^2^c = 0.8582) and time until birth (AIC = 1848.0, R^2^m = 0.0100, R^2^c = 0.8676) were representative of mHRV trends, with significantly stronger fit for weeks until birth (relative log-likelihood ratio= 859.4). This study suggests that wearable technology, such as the WHOOP strap, may provide a digital biomarker for preterm delivery by screening for changes in nighttime mHRV throughout pregnancy that could in turn alert to the need for further evaluation and intervention.

## Introduction

The United States’ singleton preterm birth rate is 8.42%, with complications related to preterm delivery resulting in increased morbidity and mortality along with an economic burden of $25.2 billion annually (Waitzman et al., 2021; Martin and Osterman, 2021). Limited screening options for the risk of preterm birth exist; those that exist, including cervical length measuring and fetal fibronectin testing, are used infrequently (Faron et al., 2020; Yost, 1999). Therefore, in practice most preterm labor has no early indications. However, when prematurity risk is identified, life and cost-saving interventions are available, most notably the use of corticosteroids and magnesium to accelerate pulmonary and nervous system development, respectively.

Heart rate variability (HRV) is a noninvasive measure of the autonomic nervous system. The WHOOP Strap (Whoop, Inc., Boston, MA, USA) is a commercially available wearable device that provides continuous physiological data including HRV for a range of applications, including predicting COVID-19 (Miller et al., 2020) and risk of adverse mental health outcomes (Czeisler et al., 2022)). Rowan et al. (2022) utilized the WHOOP Strap to describe decreases in maternal HRV during term pregnancies until approximately 33 weeks when the trend reversed and increased through delivery. All pregnancies in that study were delivered at term, leaving the cardiovascular trends that occur throughout pregnancy in preterm births unexamined. We analyzed changes in maternal HRV during pregnancy and their relation to gestational age at delivery to explore whether third trimester vital sign inflections are a feature of 33 weeks gestational age, or a feature of being seven weeks out from delivery. If inflection is predictive of time to delivery, it may provide a novel digital biomarker for prematurity.

## Materials and Methods

### Data Collection

Participants were recruited via a reproductive health survey from the existing WHOOP member base in March 2022 if they were either pregnant or had delivered between March 2021 through March 2022. Respondents who indicated in the initial survey that they were in their third trimester received a second survey in June 2022 and those who indicated that they were in their second trimester received a second survey in October 2022. Respondents who indicated in the initial survey that they were in their first trimester were not followed up with.

Participants who completed the postpartum survey, either in the initial March 2022 cohort or in the subsequent June 2022 or October 2022 cohorts were asked for their due date, delivery date, whether they had a singleton birth, and if their labor and birth were naturally initiated, as opposed to induced or scheduled for any reason. Respondent demographics including age, height, and weight were available from existing WHOOP profiles. All respondents were using a wearable device, The WHOOP Strap (Whoop, Inc., versions 3.0 or 4.0; Boston, MA, USA), which continuously collects maternal vital signs, including HRV. HRV is calculated by the WHOOP cloud-based analytics platform during non-wake periods of the primary sleep episode using the root-mean-square of successive differences method (Shaffer and Ginsberg, 2017). The WHOOP cardiovascular, respiratory, and sleep measures have been validated in a general population against gold-standard electrocardiogram and polysomnography measures and have been found to have a low degree of bias and low precision errors (Berryhill et al., 2020; Miller et al., 2022). Since data were not identifiable and were stored on a secure server, this study was deemed exempt from Institutional Review Board (IRB) oversight by Advarra’s IRB (Columbia, MD).

### Inclusion criteria

Inclusion criteria included maternal age of over 18, having an active WHOOP membership at the time of solicitation, and self-reporting of a pregnancy that resulted in live, singleton birth between March 2021 and October 2022. Participants who recorded fewer than two full nights of sleep per week from their 24th week of pregnancy through birth, gave birth to multiples, reported a birth occurring 2 or more weeks past their due date, or had scheduled inductions or cesarians were excluded from the study.

This study uses standard definitions from the American College of Obstetricians and Gynecologists and defines preterm as delivering between 20 and 37 weeks gestational age and term births as delivering between 37 and 42 weeks gestational age (American College of Gynecology, 2022). Post-term births, defined as any birth taking place at 42 or more weeks gestational age, were excluded from analysis.

### Statistical Analysis

All data were analyzed using the R programming language (version 4.2.1, Team, 2021). Modeling was performed using the stats package (version 4.3.0) and lmerTest package (version 3.1.3) (Kuznetsova et al., 2017). Data were expressed as mean ± standard deviation and differences tested by two-tailed t-test. Significance was set a priori at 0.05. Subjects were grouped into preterm and term birth subject populations. To explore the inflection point of HRV in pregnancy, for each population we fit two mixed-effect spline models to the average weekly HRV within-subject by gestational age and by weeks from birth. A spline model was selected due to the nonlinear nature of HRV during the third trimester of pregnancy (Rowan et al., 2022). A mixed-effect model was selected due to the natural variations of HRV from subject to subject.

Model 1 utilized average weekly HRVs as the response variable and gestational week starting from week 24 until the reported date of birth as the explanatory variable in a linear spline model to assess if gestational age explains changes in HRV. Based on the findings by Rowan et al., 2022, that the declining trend in HRV reverses at 33 weeks gestational age, a linear spline model with a knot at 33 gestational weeks was fit for each group and evaluated for significance and fit. Model 2 utilized average weekly HRVs as the response variable and weeks until birth from the actual date of birth backward until week 24 as the explanatory variable in a linear spline model to assess if time until birth explains changes in HRV. Based on the findings by Rowan et al., 2022, that the trend in RHR and HRV reverses 7 weeks prior to delivery, a linear spline model with a knot 7 weeks from birth was fit to each group and evaluated for significance and fit.

We compared the models within each pregnancy group using the Akaike information criterion (AIC) and R^2^. Both marginal R^2^ (R^2^m), which corresponds to variance accounted for by fixed effects, and conditional R^2^ (R^2^c), which corresponds to variance accounted for by random effects, are calculated to account for variation explained by fixed and random effects. We then compare the AIC of the two models using the relative log-likelihood ratio to quantify the difference in model performances.

## Results

### Relationship Between Maternal HRV and Gestational Age at Birth

A total of 241 individuals were eligible for analysis (**Table 1**). 21 eligible participants (8.7%) recorded preterm births, and 220 (91.3%) participants reported term pregnancies. Participants recorded an average of 99.9 (S.D. ± 19.3) days of data from their 24th week of pregnancy through birth with a total of 24,068 nighttime HRVs available for analysis. The duration of term pregnancies was on average 277.1 (S.D. ± 7.3 days) days, while the duration of preterm pregnancies was on average 246.2 (S.D ± 16.0 days) days. Participant ages ranged from 23 to 47 years (33 years, S.D ± 3.7 years). The two groups were demographically and anthropometrically similar, and failed significance testing for differences in age (P = 0.62) and BMI (P = 0.30).

**Table 1:**
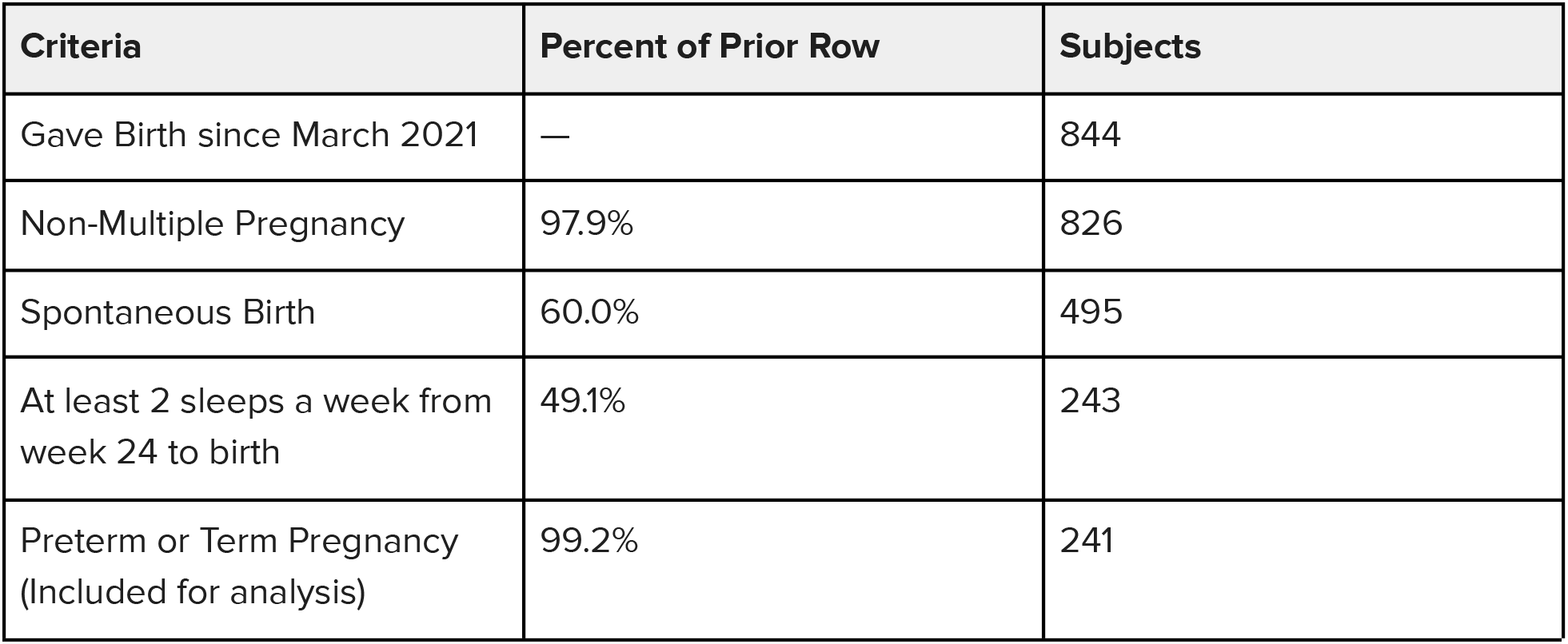
Study populations selection criteria and frequency

To determine if the rate of change in HRV is better explained by gestational age or time from birth in preterm or term pregnancies, we fit linear spline models to each group of pregnancies (**Figures 1-4)**. For term pregnancies (**Figures 1 and 2)**, gestational age (AIC = 26627.6, R^2^m = 0.0109, R^2^c= 0.8571) and weeks until birth (AIC =26616.3, R^2^m = 0.0112, R^2^c = 0.8576) explained the trends in HRV, with a stronger fit (relative log-likelihood ratio = 279.5) when aligning by weeks until birth. For preterm pregnancies (**Figures 3 and 4**), gestational age (AIC =1861.9, R^2^m = 0.0016, R^2^c = 0.8582) and time until birth (AIC = 1848.0, R^2^m = 0.0100, R^2^c = 0.8676) explained trends until birth, with a stronger fit (relative log-likelihood ratio = 1018.0) when aligning by time until birth. Differences in width of 95% confidence intervals for preterm and term pregnancies are largely explained by the significant differences in the number of subjects between the two groups.

**Figure 1:**
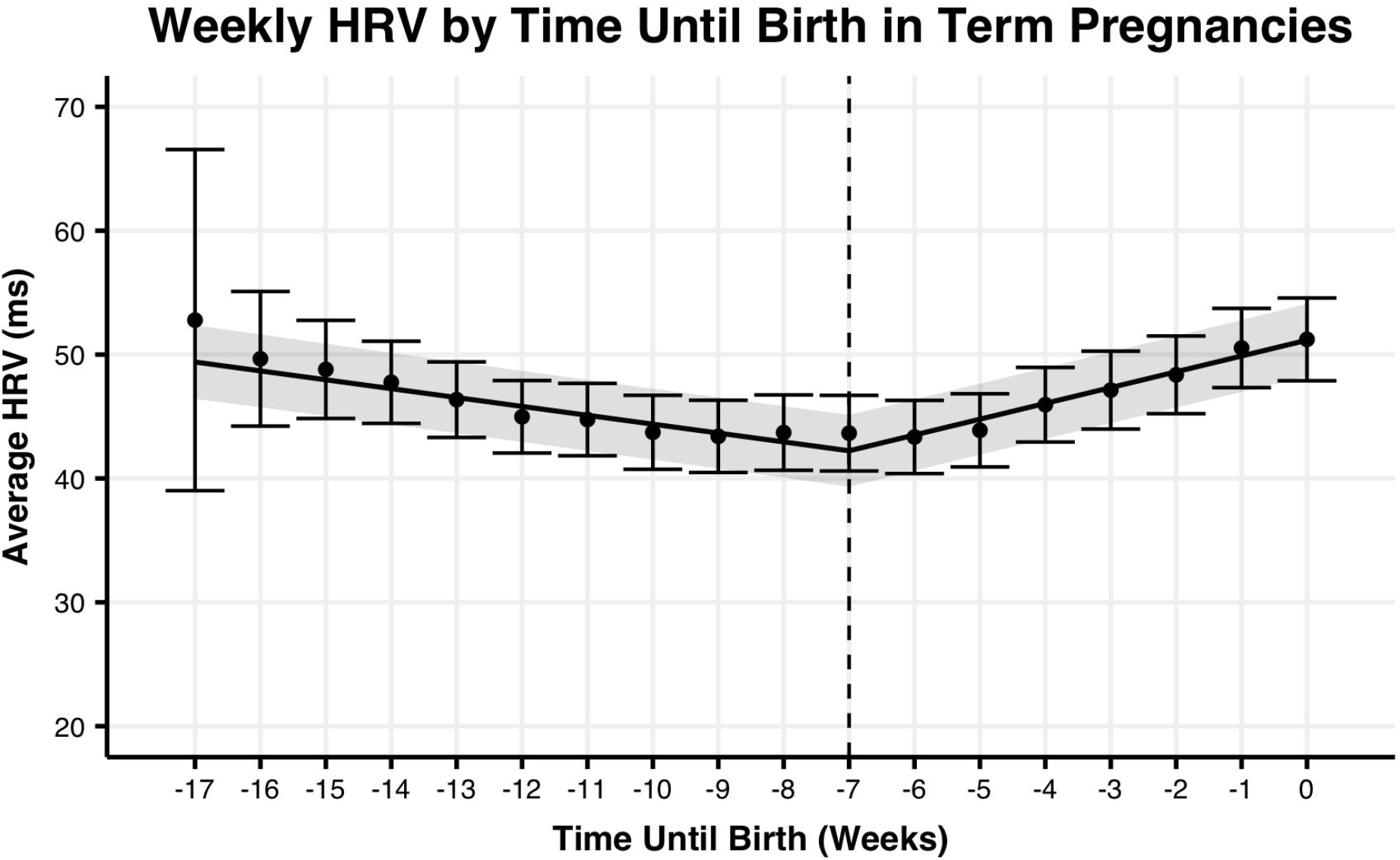
Average weekly maternal HRV from week 24 of pregnancy with to delivery date by weeks relative to delivery for term pregnancies. The trend line represents the within-person linear mixed-effects model of weekly maternal HRV, with the 95% confidence interval represented by the shaded region. Points represent the average maternal HRV by week, with the 95% confidence interval represented by the error bars.

**Figure 2:**
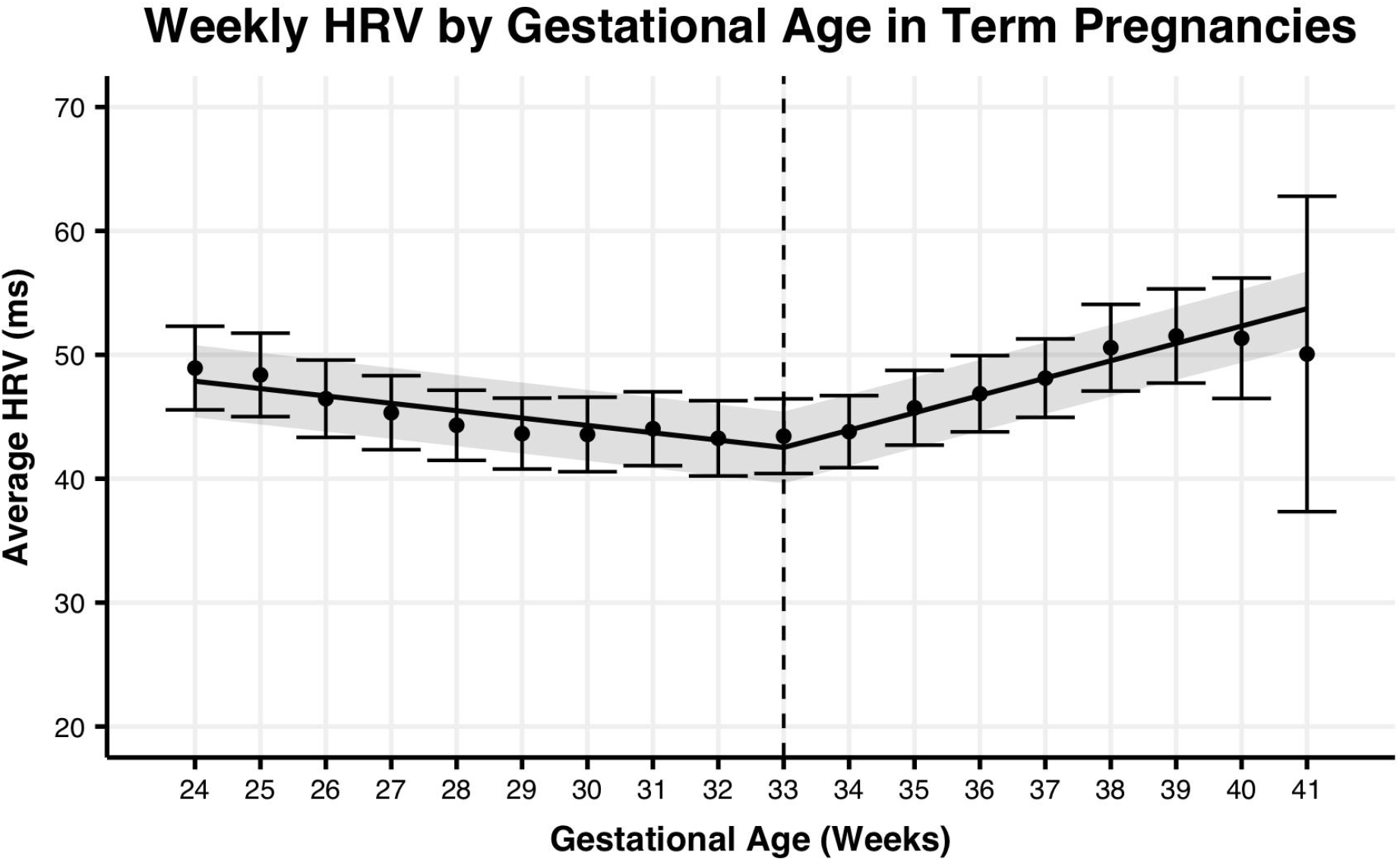
Average weekly maternal HRV from week 24 of pregnancy to delivery date by gestational age for term pregnancies. The trend line represents the within-person linear mixed-effects model of weekly maternal HRV, with the 95% confidence interval represented by the shaded region. Points represent the average maternal HRV by week, with the 95% confidence interval represented by the error bars.

**Figure 3:**
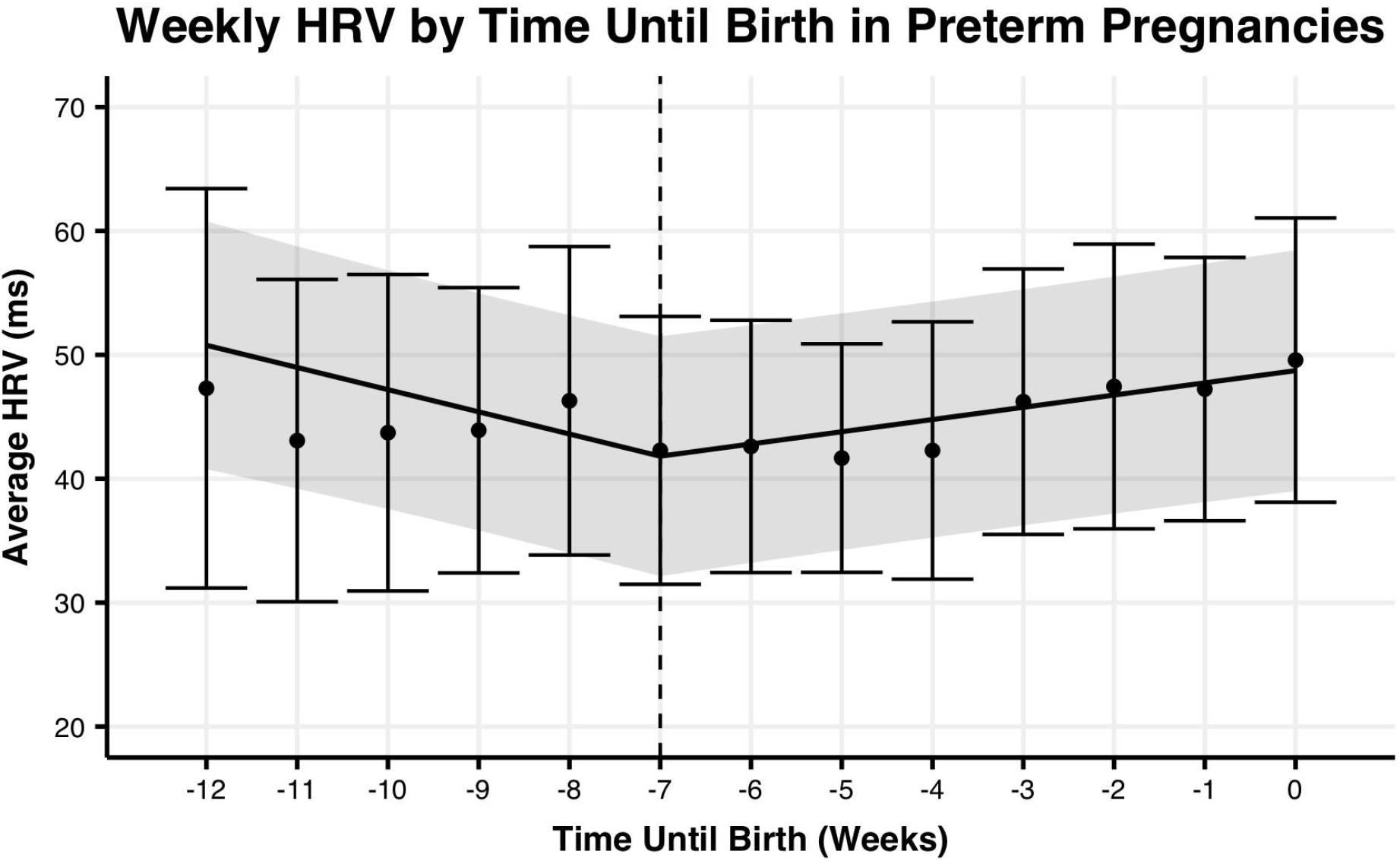
Average weekly maternal HRV from week 24 of pregnancy to delivery date by weeks relative to delivery for preterm pregnancies. The trend line represents the within-person linear mixed-effects model of weekly maternal HRV, with the 95% confidence interval represented by the shaded region. Points represent the average maternal HRV by week, with the 95% confidence interval represented by the error bars.

**Figure 4:**
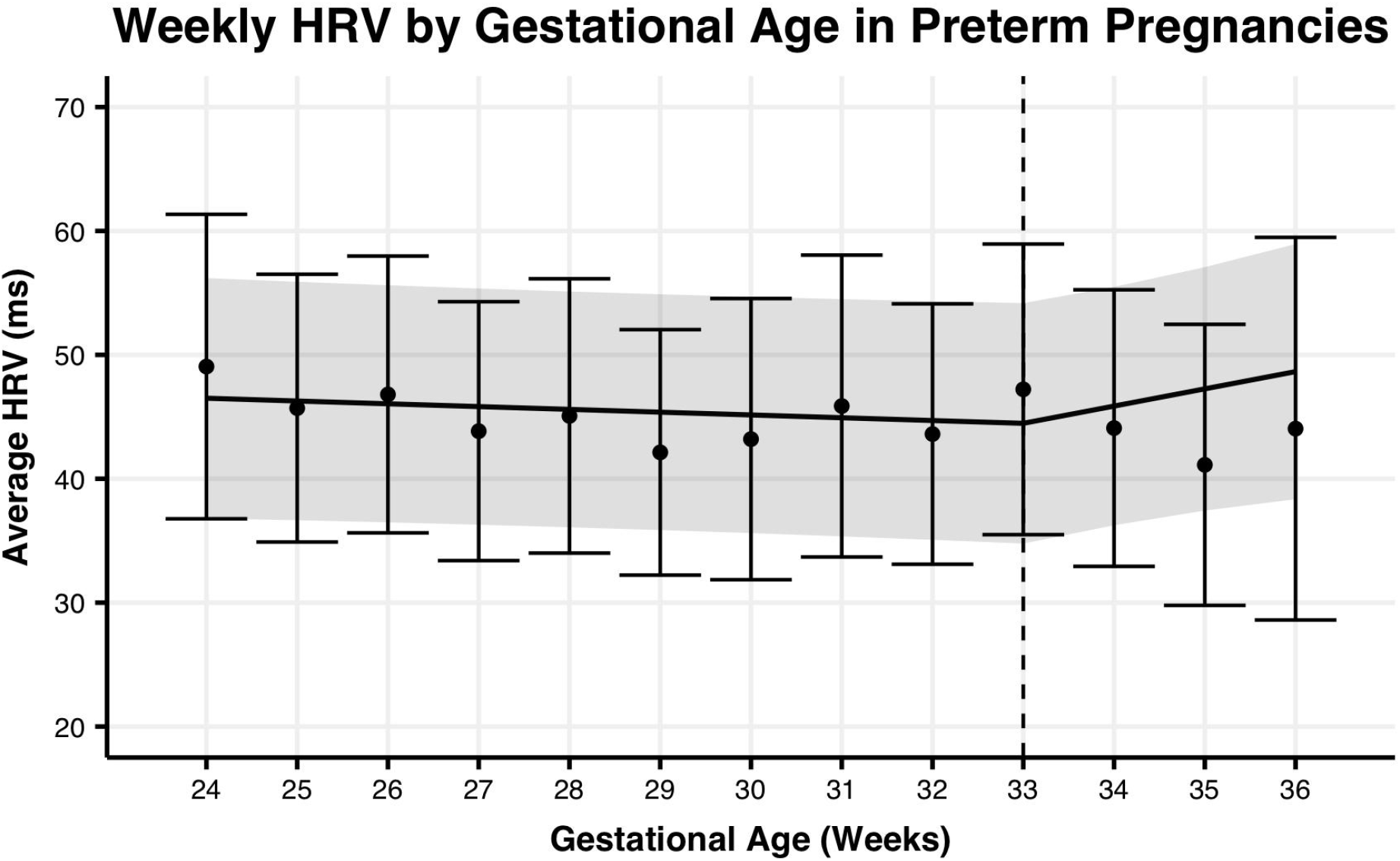
Average weekly maternal HRV from week 24 of pregnancy to delivery date by gestational age for preterm pregnancies. The trend line represents the within-person linear mixed-effects model of weekly maternal HRV, with the 95% confidence interval represented by the shaded region. Points represent the average maternal HRV by week, with the 95% confidence interval represented by the error bars.

## Discussion

Preterm delivery is difficult to predict and carries a large risk of acute and chronic morbidity, mortality, and health care costs. Transvaginal ultrasound measurements of cervical length, fetal fibronectin, and other tests for prematurity have not been widely utilized. Our study suggests that non-invasive maternal HRV monitoring may be used as a marker of time until birth in both term and preterm pregnancies based on the presence of an inflection in nightly maternal HRV. Monitoring for an early inflection in maternal HRV may provide pregnant individuals with a valuable indication that further testing for prematurity is warranted. Confirming the prematurity risk with established methods, such as fetal fibronectin testing or cervical length measuring, would allow for interventions such as progesterone to delay labor onset or antenatal corticosteroid and magnesium administration, which could respectively contribute to improved respiratory and nervous system function at birth, reducing time spent in the neonatal intensive care unit and resulting in long-term better health outcomes.

As more women find themselves in obstetric deserts, detecting the timing of preterm and term deliveries can help them be more prepared and enable them to access a properly equipped delivery facility in a timely fashion. There are substantial racial and ethnic disparities in preterm rates and obstetric deserts, with those in historically-minoritized populations having significantly higher preterm rates than white populations (Martin and Osterman, 2021), thereby allowing these interventions to offset these inequities.

In a 2019 online survey, 78% of respondents reported a willingness to continuously wear a device to monitor maternal and fetal wellbeing during pregnancy (Wakefield et al., 2022). This finding suggests that non-invasive wearable technology, such as the WHOOP strap, offers an acceptable way to intervene in potential preterm delivery.

### Strengths and Limitations

There are several strengths of this study. Our sample size is larger than previous studies and especially with the continuous data points, allows for meaningful conclusions to be drawn. Additionally, we reproduced the results of the prospective study conducted by Rowan et al. (2022) with women recruited from a single site fertility clinic in West Virginia experiencing singleton pregnancies. Our study represents women from 42 US states and territories and 16 countries.

The subjects in this study were active members of the WHOOP platform, and therefore may not be nationally representative when it comes to socioeconomic factors, activity levels, or access to healthcare. Nonetheless, our observed rate of preterm delivery was 8.7%, which is consistent with the national average of 8.42% (Martin and Osterman, 2021).

This study relied on retrospective survey data collected postpartum via an in-app survey and is therefore subject to human error in survey response data. Race, ethnicity, and socioeconomic status were not collected in this study. The survey was administered to mothers following delivery of live babies and therefore did not include women whose pregnancies did not result in a live birth. Further research is required to understand the relationship between maternal HRV and birth outcomes other than live birth, including stillbirth and miscarriage. The current dataset is underpowered to explore the predictive odds of early vital sign inflection as an indicator of preterm birth at the individual level, additional research at a larger scale is warranted to explore this further.

### Conclusion and Future Directions

This study is the first to demonstrate that maternal HRV during pregnancy differs in preterm and term births in singleton pregnancies. Wrist-worn, noninvasive wearable devices present an exciting opportunity for monitoring, allowing for continuous tracking of health metrics such as maternal HRV. The ability for continuous passive monitoring becomes especially important for pregnancies in medically underserved areas and obstetric deserts where prematurity is known to have comparatively worse outcomes than areas with abundant obstetric resources. Regardless of proximity to obstetric care, since prematurity risk isn’t routinely screened for due to limitations in extant methodologies (Faron et al., 2020; Yost, 1999), many women who would benefit from interventions such as antenatal corticosteroids and magnesium are not administered these medications in a time frame that would allow for productive intervention. The screening provided by the passive continuous monitoring of wearable devices could allow for early detection of changes in HRV, potentially alerting to increased prematurity risk.

Future prospective studies are needed to determine whether preterm delivery can be predicted in real time at the individual level using wearable technology. There is increasing evidence that physical activity and nutrition-based interventions targeting modifiable risk factors could reduce the risk of preterm delivery (Thoene et al., 2020; Catov et al., 2018). HRV can also be improved with changes to sleep, exercise, and nutrition (Routledge et al., 2010; Sajjadieh et al., 2020; Young and Benton, 2018). Therefore, interventional studies that modify HRV are warranted to explore the potential to manipulate the timing of the HRV inflection in order to reduce prematurity. Together, the future directions and implications of this study allow for greater understanding and intervention of pregnant individuals who face preterm delivery.

## Data Availability

Add data produced in the present study are protected intellectual property of WHOOP, Inc., however may be made available upon request for researchers who meet the criteria for access to confidential information.

## Author Contributions

ERC and SRJ designed the study and conducted the analysis. DMP provided statistical and analytical support. SR aided in the medical interpretation of the findings. EAC assisted in literature review and contextualizing findings within the literature. All authors contributed to the writing and editing of the manuscript.

## Additional Acknowledgements

The authors would like to recognize the contributions of Laura Ware who participated in workshopping the analytical approaches explored in preparing this manuscript.

## Author Access to Data

Only ERC, SRJ, and DMP had access to the data analyzed in this manuscript.

